# COVID-19 vaccine effectiveness among immunocompromised populations: a targeted literature review of real-world studies

**DOI:** 10.1101/2021.12.29.21268511

**Authors:** Manuela Di Fusco, Jay Lin, Shailja Vaghela, Melissa Lingohr-Smith, Jennifer L Nguyen, Thomas Scassellati Sforzolini, Jennifer Judy, Alejandro Cane, Mary M Moran

## Abstract

**Introduction:** From July through October of 2021, several countries issued recommendations for increased COVID-19 vaccine protection for individuals with one or more immunocompromised (IC) conditions. It is critically important to understand the vaccine effectiveness (VE) of COVID-19 vaccines among IC populations as recommendations are updated over time in response to the evolving COVID-19 pandemic.

**Areas covered:** A targeted literature review was conducted to identify real-world studies that assessed COVID-19 VE in IC populations between December 2020 and September 2021. A total of 10 studies from four countries were identified and summarized in this review.

**Expert opinion/commentary:** VE of the widely available COVID-19 vaccines, including BNT162b2 (Pfizer/BioNTech), mRNA-1273 (Moderna), Ad26.COV2.S (Janssen), and ChAdOx1 nCoV-19 (Oxford/AstraZeneca), ranged from 64%-90% against SARS-CoV-2 infection, 73%-84% against symptomatic illness, 70%-100% against severe illness, and 63%-100% against COVID-19-related hospitalization among the fully vaccinated IC populations included in the studies. COVID-19 VE for most outcomes in the IC populations included in these studies was lower than in the general populations. These findings provide preliminary evidence that the IC population requires greater protective measures to prevent COVID-19 infection and associated illness, hence should be prioritized while implementing recommendations of additional COVID-19 vaccine doses.

## 1. Introduction

As of September 30, 2021, approximately 45% of the worldwide population had received at least one dose of a coronavirus disease 2019 (COVID-19) vaccine [1]. Scientific evidence gained from real-world studies conducted in multiple countries is increasingly showing that widely available COVID-19 vaccines, including BNT162b2 (Pfizer/BioNTech), mRNA-1273 (Moderna), Ad26.COV2.S (Janssen), and ChAdOx1 nCoV-19 (Oxford/AstraZeneca), are effective against severe acute respiratory syndrome coronavirus 2 (SARS-CoV-2) infection, symptomatic COVID-19 illness, and COVID-19-related hospitalization and death [2,3]. Such findings from real-world studies are generally consistent with the efficacy results of the randomized clinical trials (RCTs) of these vaccines [4-7]. Vaccine efficacy in clinical trials and vaccine effectiveness (VE) measured in real-world studies both calculate the risk of disease among vaccinated and unvaccinated individuals and the percentage reduction in risk of disease among vaccinated individuals relative to unvaccinated individuals; VE equates to the reduction in disease occurrence for those who are vaccinated (i.e., a VE of 85% = an 85% reduction in disease occurrence among the vaccinated) [8].

From July through October of 2021, several countries across the world issued recommendations for increased COVID-19 vaccine protection for individuals with one or more immunocompromised (IC) conditions; many of these recommendations also included other subpopulations (e.g., elderly) [9]. IC individuals are generally defined as those with suppressed immunity resulting from health conditions (e.g., organ transplant, malignancy, rheumatological condition, human immunodeficiency virus [HIV] infection, etc.) and/or with active usage of immunosuppressive medications [10]. These recommendations were informed by real-world studies of IC populations, who were largely excluded from the RCTs of the COVID-19 vaccines [4-7], that observed a reduced immune response to COVID-19 vaccines in IC individuals compared to the general population [11-16]. Whether a reduced immune response to COVID-19 vaccines correlates with diminished VE is not well understood. However, results from several recent real-world studies conducted in the United States (US), Israel, England, and Qatar that assessed VE in IC populations [17-27] indicate that IC individuals are at increased risk for severe COVID-19 outcomes. These studies found that although the COVID-19 vaccines provided a high level of protection against SARS-CoV-2 infection, symptomatic COVID-19 illness, and/or COVID-19-related hospitalization, VE for the IC populations tended to be lower than that observed in the general population [17-27].

As new COVID-19 vaccine recommendations are implemented and updated over time in response to the evolving COVID-19 pandemic, it is necessary to rapidly and more comprehensively understand the effectiveness of COVID-19 vaccines in IC populations. From a policy perspective, such information could provide decision makers with the data to help to fill vaccine coverage gaps and instill greater protective measures towards the IC population, measures such as additional dose/booster prioritization. This objective has become even more critical given the continuing risk of emergence of more transmissible variants (i.e., Omicron). Towards, this objective, in this review, we have summarized the findings of real-world studies that have assessed COVID-19 VE in IC populations.

## 2. Methods

### 2.1. Research question and study inclusion criteria

The research question and study eligibility criteria were developed based on the Population, Intervention, Comparator, Outcomes (PICO) framework [28]. The research question was, what is the reported COVID-19 VE in IC populationsã IC populations were defined according to the definitions used in the individual studies. The interventions assessed were any of the widely available COVID-19 vaccines in the world. The outcomes explored included COVID-19 VE against SARS-CoV-2 infection, symptomatic COVID-19 illness, severe COVID-19 illness, and COVID-19-related hospitalization/death. We targeted real-world observational studies, either cross-sectional or longitudinal in design, conducted in any country that assessed these outcomes and reported calculated VE estimates. Studies that evaluated vaccine efficacy in the context of a clinical trial or immunogenicity were not included in this review.

### 2.2. Search strategy and screening

The best practice in systematic literature reviews is to prioritize searches and to include studies that are peer reviewed and published [29]. Given that the interventions (i.e, COVID-19 vaccines), in the scope of this review were recently introduced, and that there has been a high influx of COVID-19 research being posted on pre-print servers, we included both peer-reviewed and non-peer-reviewed preprint studies. While this approach strengthens the comprehensiveness of this review, the author team recognizes the potential limitations in the reproducibility of the review and the quality of the collected evidence base.

Based on the above, a targeted search was performed using PubMed and the preprint servers, medRxiv and Khub, to identify real-world studies that assessed COVID-19 VE in IC populations between December 2020 and September 30, 2021 (inclusive). The following list of terms was generated and searched across all study fields: “COVID-19”, “SARS-CoV-2”, “vaccine effectiveness”, and “immunocompromised”. To maximize the scope of the search, no search terms were included for interventions or outcomes. All studies found written in the English language, without restrictions of countries, but with reported COVID-19 VE against SARS-CoV-2 infection, symptomatic COVID-19 illness, severe COVID-19 illness, and/or COVID-19-related hospitalization/death were examined for inclusion. Titles, abstracts, and full study contents publicly available were screened by one independent reviewer (MLS). Since there was only one reviewer, random selection and inter-rater reliability scores (e.g., kappa) were not determined.

### 2.3. Data extraction

Study characteristics (i.e., countries, vaccines included in analyses, study periods, study designs, and data sources), general characteristics of the overall study population (i.e., sample size, follow-up duration, proportion of fully vaccinated, median age, and sex distribution), IC definitions and IC population characteristics, and details of the study outcome measures related to VE (i.e., controls, VE follow-up durations, VE calculations, and analysis methods) were extracted and incorporated into an Excel spreadsheet. When possible, COVID-19 VE in the general and non-IC population was also extracted. Since all data presented in this review were extracted from already published and/or publicly available preprint studies, this review is not subject to ethical approval.

### 2.4. Narrative synthesis

Given the diversity of the studies included, the quality of the selected studies was not compared, and meta-analyses were not performed. The assembled body of evidence was drawn together and interpreted in a narrative synthesis. After tabulating the individual studies, we assessed if the observed outcomes of interest were consistent across studies, which were interpreted in the context of their similarities (e.g., definition of symptomatic COVID-19 illness) and differences (e.g., VE follow-up durations). We qualitatively grouped the studies by outcome measures and investigated any reasons for inconsistencies among the results. This approach is supported by guidance for undertaking reviews [30].

## 3. Results

### 3.1. Search results

With an end search date of September 30, 2021, a total of 10 studies were identified in which COVID-19 VE was assessed in IC populations [17-27]; six, with one study accounting for two publications, were peer-reviewed [17,18,19,20,22,23,24] and four were preprints [21,25,26,27] at the time this review was written. The study outcomes included COVID-19 VE against SARS-CoV-2 infection, symptomatic COVID-19 illness, severe COVID-19 illness, and COVID-19-related hospitalization, which were summarized for the study IC populations, as well as the general populations, when such data were available. Although one study assessed COVID-19 VE against all-cause death [20], VE specifically against COVID-19-related death was not reported in the included studies for IC populations and therefore was not summarized in this review.

### 3.2. Key study characteristics

Tables 1, 2, and 3 show study characteristics, general characteristics of the overall study populations, and a comparison of IC definitions and IC populations across studies, respectively.

**Table 1.** Study characteristics.

| **Study/Peer-reviewed** | **Country** | **Vaccines included in analyses** | **Study period** | **Study design** | **Data source** |
| --- | --- | --- | --- | --- | --- |
| Young-Xu Y, et al./Yes [17] | United States | • Pfizer-BioNTech BNT162b2 mRNA vaccine<br>• Moderna mRNA-1273 vaccine | Pre-Delta:<br>Dec 14, 2020 – Mar 14, 2021 | • Matched test negative case-control analysis for infection<br>• Matched case-control analysis for hospitalization and death | EMR data from VHA Corporate Data Warehouse |
| Tenforde MW, et al./Yes [18] | United States | • Pfizer-BioNTech BNT162b2 mRNA vaccine<br>• Moderna mRNA-1273 vaccine | Intermediate:<br>Mar 11 – May 5, 2021 | Case-control analysis | Hospital admission logs and EMRs (18 academic hospitals/16 states) |
| Tenforde MW, et al./Yes [19] | United States | • Pfizer-BioNTech BNT162b2 mRNA vaccine<br>• Moderna mRNA-1273 vaccine | Including Delta:<br>Mar 11 – Jul 14, 2021 | Case-control analysis | Hospital admission logs and EMRs (21 academic hospitals/18 states) |
| Khan N & Mahmud N/Yes [20] | United States | • Pfizer-BioNTech BNT162b2 mRNA vaccine<br>• Moderna mRNA-1273 vaccine | Pre-Delta:<br>Dec 18, 2020 – Apr 20, 2021 | Retrospective cohort analysis | VHA data sources, including VHA Corporate Data Warehouse |
| Polinski JM, et al./No, medRxiv preprint [21] | United States | • Janssen Ad26.COV2.S vaccine | Including Delta:<br>Mar 1 – Jul 31, 2021 | Matched control analysis | Administrative insurance claims in Health Verity database |
| Dagan N, et al.; Barda N, et al./Yes [22,23] | Israel | • Pfizer-BioNTech BNT162b2 vaccine | Pre-Delta:<br>Dec 20, 2020 – Feb 1, 2021 | Matched control analysis | EMR data from Clalit Health Services data repositories |
| Chodick G, et al./Yes [24] | Israel | • Pfizer-BioNTech BNT162b2 vaccine | Pre-Delta:<br>Dec 19, 2020 – Feb 20, 2021 | Retrospective cohort analysis | EMR data from Maccabi Healthcare Services databases |
| Yelin I, et al./No, medRxiv preprint [25] | Israel | • Pfizer-BioNTech BNT162b2 vaccine | Pre-Delta:<br>Dec 1, 2020 – Feb 25, 2021 | Prospective patient-level analysis | EMR data from Maccabi Healthcare Services databases |
| Whitaker HJ, et al./No, Khub preprint [26] | England | • Pfizer-BioNTech BNT162b2 mRNA vaccine<br>• Oxford/AstraZeneca ChAdOx1 nCoV-19 vaccine | Including Delta:<br>Dec 7, 2020 – Jun 13, 2021 | Nested test-negative case-control analysis | EMR data of 718 general practices |
| Chemaitelly H, et al./No, medRxiv preprint [27] | Qatar | • Pfizer-BioNTech BNT162b2 mRNA vaccine<br>• Moderna mRNA-1273 vaccine | Including Delta<sup>a</sup>:<br>Feb 1, 2021 – Jul 21, 2021 | Retrospective cohort analysis with cross-over design | Hamad Medical Corporation: integrated nationwide digital-health information platform |

EMR: Electronic medical record; mRNA: messenger ribonucleic acid; VHA: Veterans Health Administration
Covariant predominance period was defined as Pre-Delta, Intermediate, and Including Delta (when the study period extended to months when the Delta variant was predominant) for countries referring to https://covariants.org/per-country, as below:
**US:** Dec 2020-April 2021 as Pre-delta; May 2021-June 2021: Intermediate; July 2021-Sept 2021: Delta.
**Israel:** Dec 2020 – Feb 2021 was Alpha, which was defined as Pre-delta.
**England:** Dec 2020 – April 2021 was Alpha (Pre-delta), May 2021: Intermediate, June 2021 was during Delta variant, hence defined as including Delta.
**Qatar:** Feb-Apr 2021 was Beta (Pre-delta), May 2021: Intermediate, and June-July 2021 was Delta, hence as defined as including Delta.
<sup>a</sup> In Qatar, the Delta variant was preceded by the Beta variant, as opposed to the Alpha variant in the US, Israel, and England. Authors reported that as of July 28, 2021, Delta was at low incidence in Qatar.

**Table 2.** General characteristics of the overall study populations.

| **Study** | **Count** | **Follow-up duration** | **Proportion fully vaccinated** | **Age** | **Sex distribution** |
| --- | --- | --- | --- | --- | --- |
| Young-Xu Y, et al. [17] | Overall: N=6,647,733 in VHA cohort<br>SARS-CoV-2 positive cases: N=15,110<br>Controls: N=60,436 | ≥14 days after 2<sup>nd</sup> dose up to 3 months | Positive cases: N=41 (0%)<br>Controls: N=3,711 (6%) | Median age not reported.<br>Positive cases: 66% were 45-74 yrs of age<br>Controls: 78% were 45-74 yrs of age | Positive cases: 90% male<br>Controls: 90% male |
| Tenforde MW, et al. [18] | Overall: N=1,212 in general hospitalized population<br>Hospitalized COVID-19 cases: N=593<br>Controls: N=619 | Median: 43 days; maximum: 113 days | Cases: N=45 (8%)<br>Controls: N=215 (35%) | Median age of cases: 56 yrs<br>Median age of controls: 61 yrs | Cases: 48% female<br>Controls: 51% female |
| Tenforde MW, et al. [19] | Overall: N=3,089 in general hospitalized population<br>Hospitalized COVID-19 cases: N=1,194<br>Controls: N=1,895 | Up to 24 weeks | Cases: N=141 (12%)<br>Controls: N=988 (52%) | Median age of cases: 56 yrs<br>Median age of controls: 62 yrs | Cases: 49% female<br>Controls: 49% female |
| Khan N & Mahmud N. [20] | Overall: N=14,697 in VHA cohort with IBD<br>Vaccinated: N=7,321<br>Unvaccinated: N=7,376 | Vaccinated: median of 38 days<br>Unvaccinated: median of 123 days | 91% of Pfizer (N=3,017) and 89% of Moderna (N=3,561) patients received both vaccine doses during full study period | Median age of vaccinated: 71 yrs<br>Median age of unvaccinated: 64 yrs | Vaccinated: 93% male<br>Unvaccinated: 92% male |
| Polinski JM, et al. [21] | General population cohort<br>• Vaccinated: N=390,517<br>• Unvaccinated: N=1,524,153 | ≥14 days after single dose and up to 5 months | 100% of vaccinated cohort | Median age of vaccinated: 55 yrs<br>Median age of unvaccinated: 55 yrs | Vaccinated: 56% female<br>Unvaccinated: 56% female |
| Dagan N, et al.; Barda N, et al. [22,23] | General population cohort<br>• Vaccinated: N=596,618<br>• Unvaccinated: N=596,618 | Mean follow-up: 15 days<br>Subgroup analyses: mean follow-up 18 days | Among those with ≥21 days of follow-up, 96% received a 2nd dose | Median age of vaccinated: 45 yrs<br>Median age of unvaccinated: 45 yrs | Vaccinated: 50% male<br>Unvaccinated: 50% male |
| Chodick G, et al. [24] | Overall: N=1,178,597 in general population<br>• Protection period group: N=872,454<br>• Reference period group: N=1,178,597 | Days 1-7 after 1<sup>st</sup> dose<br>Days 7-27 after 2<sup>nd</sup> dose | N=872,454 (74%) | Median age of protection period group: 52 yrs<br>Median age of reference period group: 48 yrs | Protection period group: 48% male<br>Reference period group: 48% male |
| Yelin I, et al. [25] | Overall in general population: 1.8 million/ 1.3 million were vaccinated | 67 days, >93 million observations after exclusion | >98% of patients administered with the 1<sup>st</sup> dose administered with 2<sup>nd</sup> dose; 1,721,377 vaccinated | Not included in preprint | Not included in preprint |
| Whitaker HJ, et al. [26] | Overall: N=5,642,687 in general population<br>Clinical risk groups: N=1,054,510 | ≥14 days after 2<sup>nd</sup> dose up to 6.5 months | Data not available; to be published in Supplementary material S2. | Data not available; to be published in Supplementary material S2. | Data not available; to be published in Supplementary material S2. |
| Chemaitelly H, et al. [27] | N=782 kidney transplant recipients<br>• Vaccinated ≥14 days after 2<sup>nd</sup> dose through study period: N=506; 248<br>crossed over from<br>unvaccinated cohort<br>• Unvaccinated:<br>N=423 | ≥14 days after 2<sup>nd</sup> dose up to ~6 months | N=601 (77% of 782 with ≥1<sup>st</sup> dose) | Median age of vaccinated: 52 yrs<br>Median age of unvaccinated: 49 yrs | Vaccinated: 70% male<br>Unvaccinated: 63% male |

COVID-19: Coronavirus disease 2019; IBD: Inflammatory bowel disease; SARS-CoV-2: Severe acute respiratory syndrome coronavirus 2; Yrs: Years; VHA: Veterans Health Administration

**Table 3.**
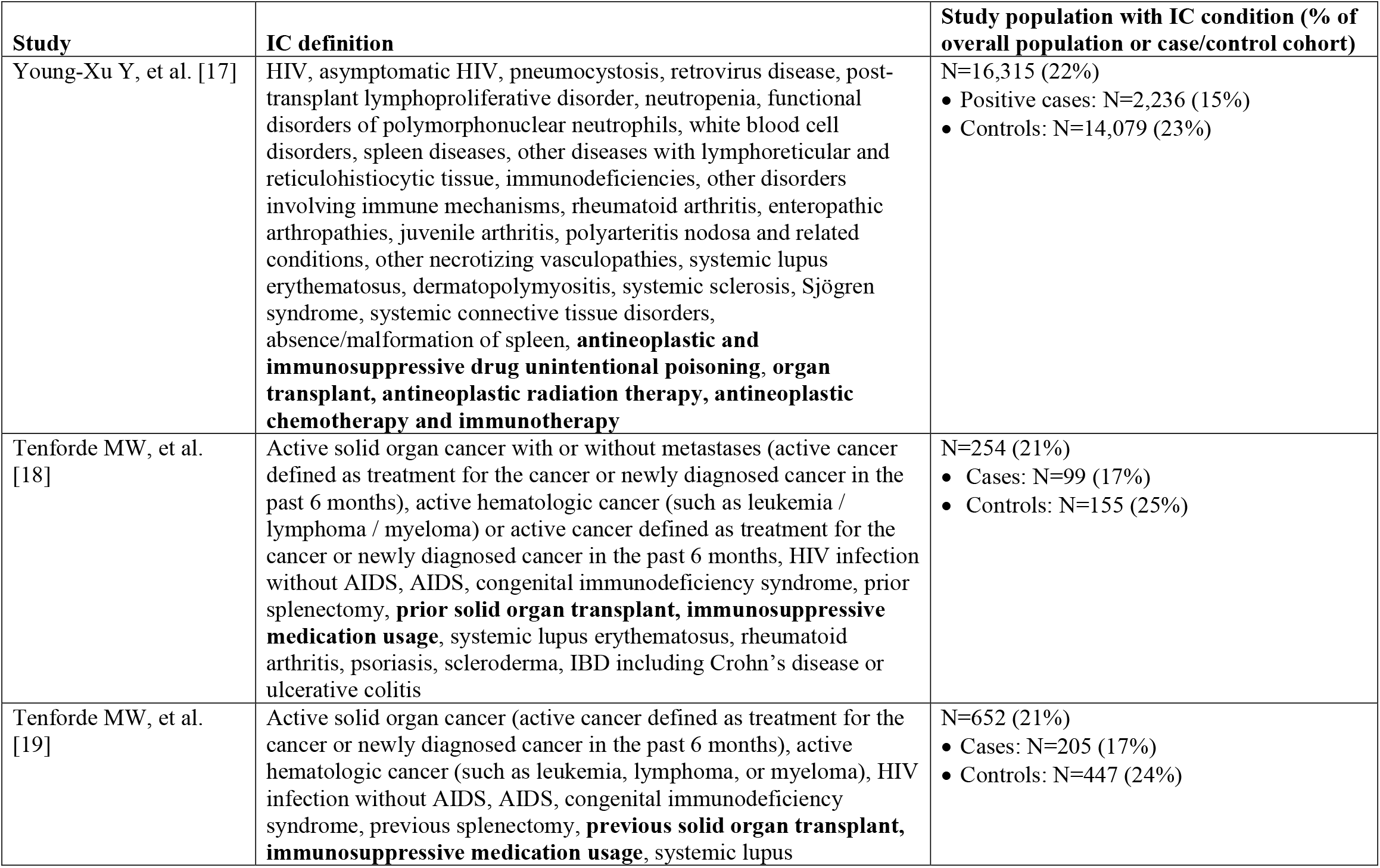
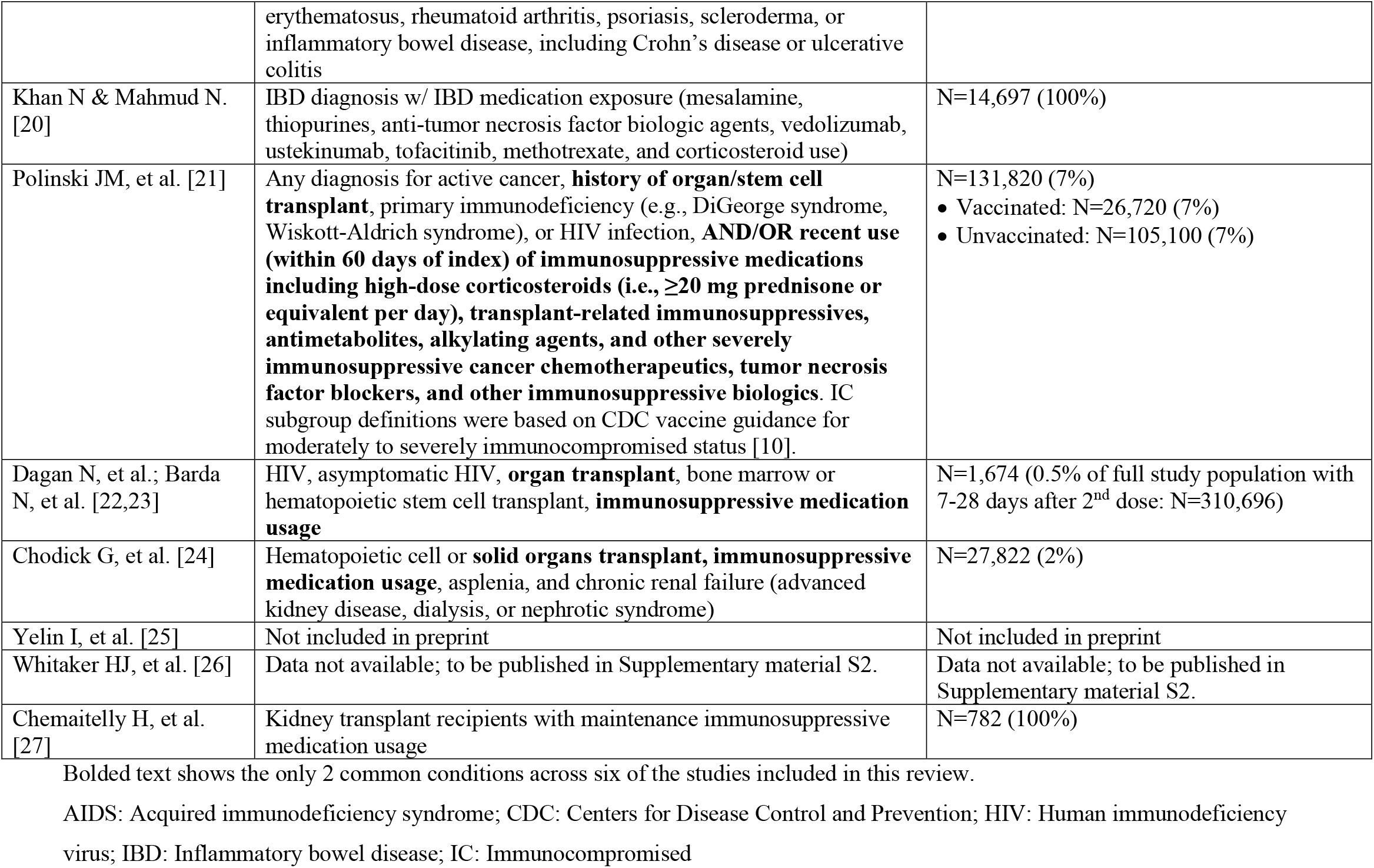
Comparison of IC definitions and populations across studies.

#### 3.2.1 Study geographical location and design

The 10 studies summarized in this review were conducted in four countries, the US, Israel, England, and Qatar. Four of the five studies conducted in the US assessed VE of the mRNA vaccines, BNT162b2 and mRNA-1273, using electronic medical records (EMRs) and/or hospital admission logs. Young-Xu et al. [17] conducted a matched analysis of COVID-19 cases and controls among US veterans with an IC subgroup; the CDC COVID-19 Response Team conducted two unmatched hospitalized COVID-19 case-control analyses among the general hospitalized population with IC subgroups [18,19], and Khan & Mahmud [20] conducted a retrospective cohort analysis of US veterans with inflammatory bowel disease (IBD). The fifth US study, by Polinski et al. [21], performed a matched control analysis of Ad26.COV2.S VE among the general population and an IC subgroup using the administrative insurance claims database of Health Verity.

Three studies, all of which assessed VE of the BNT162b2 mRNA vaccine in the general population and IC subgroups, were conducted in Israel [22,23,24,25], including the largest matched population analysis of vaccinated and unvaccinated persons (N=596,618 matched pairs) conducted to date by Dagan et al. [22], with follow-up subgroup analyses, which included an IC population, provided in Barda et al. [23]; the data source of this study was EMRs in Clalit Health Services data repositories. Chodick et al. [24] conducted a retrospective cohort analysis and Yelin et al. [25] conducted a prospective patient-level analysis; both studies extracted data from EMRs in Maccabi Healthcare Services databases.

The nested test-negative case-control study conducted in England by Whitaker et al. [26] was the only study that evaluated ChAdOx1 nCoV-19 VE in the general population with an IC subgroup. This study also assessed VE of the BNT162b2 mRNA vaccine; it utilized EMR data from 718 general practices [26]. Chemaitelly et al. [27] conducted a retrospective cohort analysis with a cross-over design of mRNA (BNT162b2 and mRNA-1273 vaccine) VE in a population of kidney transplant recipients in Qatar; data were extracted from an integrated nationwide digital-health information platform from the Hamad Medical Corporation.

The study periods (i.e., follow-up) were all shorter than 6.5 months. Five of the included studies in this review assessed COVID-19 VE primarily from December 2020 through February/March 2021 [17,20,22,23,24,25]; the three studies conducted in Israel fall within this group. Tenforde et al. [19], Polinski et al. [21], Whitaker et al. [26], and Chemaitelly et al. [27] had study periods that extended into the summer months (up until July) of 2021. Only Tenforde et al. [19] and Polinski et al. [21] separated their VE analyses by time periods to better understand if VE was affected by the emergence of the Delta variant in the US.

#### 3.2.2 Characteristics of the study IC populations

The health conditions used to define IC populations varied across the studies. In two studies, Yelin et al. [25] and Whitaker et al. [26], the definitions of IC were not available in the preprint materials. Khan & Mahmud [20] and Chemaitelly et al. [27] assessed COVID-19 VE in specific IC populations, IBD patients among US veterans and kidney transplant recipients, respectively; in both studies, patients also had maintenance immunosuppressive medication usage. In the other six studies [17,18,19,21,22,23,24], IC populations were defined according to various IC conditions; only two IC conditions, organ transplant and immunosuppressive medication usage, were common across the six studies. Other IC conditions common across multiple studies included HIV infection in five studies, active cancer in four, immunodeficiencies in four, rheumatoid arthritis/other related inflammatory conditions in three; chronic kidney disease (CKD) was included in only one study. In these six studies, some patient groups with other IC diseases that were not specifically defined may have been captured among those grouped with immunosuppressive medication usage.

The sample sizes of the IC populations were reported in eight studies and are summarized in Table 3. Sample sizes included 16,315 (22% of overall study population) in Young-Xu et al. [17], 254 (21% of overall study population) in Tenforde et al. [18], 652 (21% of overall study population) in Tenforde et al. [19], 14,697 (100% IBD population) in Khan & Mahmud [20], 131,820 (7% of overall study population) in Polinski et al. [21], 1,674 (0.5% of overall study population) in Dagan et al.; Barda et al. [22,23], 27,822 (2% of overall study population) in Chodick et al. [24], and 782 (100% kidney transplant recipients) in Chemaitelly et al. [27]. No information on sample size was available for Yelin et al. [25] and Whitaker et al. [26] at the time of writing this review.

Only three studies provided characteristics of the IC populations in which COVID-19 VE was assessed [20,21,27]. Khan & Mahmud [20] conducted their study specifically among Veterans Health Administration (VHA) patients with IBD who took immunosuppressive medications; median age was 68 years, 92% were male, 80% were White, approximately 44% were from the South US region, and 62% had ulcerative colitis. The frequency of breakthrough infections was 0.11% (N=7) in those who were fully vaccinated compared to 1.34% (N=197) among those who were not vaccinated [20]. Polinski et al. [21] defined their IC population according to the guidance of the US Centers for Disease Control and Prevention (CDC) for moderately to severely IC status [10]. In this study, the IC represented 6.8% (N=26,720) of the overall vaccinated population and 6.9% (N=105,100) of the overall unvaccinated population; mean age of matched vaccinated and unvaccinated IC groups was 59 years, 60% were female, ethnicity/race was not reported, and approximately 41% resided in the South US region [21]. Chemaitelly et al. [27] conducted their study specifically in kidney transplant recipients who were on maintenance immunosuppressive medication; the study population (N=782) was in Qatar; median age of the vaccinated cohort was 52 years and 70% were male; the median age of the unvaccinated cohort was 49 years and 63% were male [27]. The incidence of breakthrough infections was 2.58% in those who were vaccinated compared to 4.74% among those who were unvaccinated (follow-up: 120 days after 14 days after 2^nd^ dose) [27].

### 3.3. COVID-19 VE

Table 4 reports the details of the study outcome measures related to COVID-19 VE (i.e., outcome measures, controls, VE follow-up duration, VE calculations, and analysis methods), while Table 5 presents the reported VE estimates, including 95% confidence intervals (95% CI), against SARS-CoV-2 infection, symptomatic COVID-19 illness, severe COVID-19 illness, and COVID-19-related hospitalization across the studies included in this review. Figure 1 graphically presents COVID-19 VE in IC populations relative to overall study populations from those studies with such available data.

**Table 4.** Details of the study outcome measures related to COVID-19 VE.

| **Study** | **Outcome measures** | **Controls** | **VE follow-up duration** | **VE calculation** | **Analysis method** |
| --- | --- | --- | --- | --- | --- |
| Young-Xu Y, et al. [17] | • VE: SARS-CoV-2 infection (positive RT-PCR or antigen test)<br>• VE: COVID-19-related hospitalization (admission/discharge diagnosis of COVID-19); not assessed in IC<br>• VE: COVID-19-related death; not assessed in IC | • For SARS-CoV-2 infection, controls were patients with a negative SARS-CoV-2 RT-PCR test<br>• For endpoints 2 & 3, controls were patients with a positive SARS-CoV-2 RT-PCR test who were not hospitalized for COVID-19 or died during the study period | ≥14 days after 2<sup>nd</sup> dose up to 3 months | (1–OR) X 100<br>OR: SARS-CoV-2 infection in vaccinated vs. unvaccinated | Logistic regression |
| Tenforde MW, et al. [18] | • VE: Odds of prior SARS-CoV-2 vaccination in hospitalized cases with COVID-19 (symptoms and a positive RT-PCR/clinical test) vs. controls | • Test-negative controls: Hospitalized patients who tested negative for SARS-CoV-2, but with acute respiratory illness (ARI) symptoms<br>• ARI syndrome-negative controls: Hospitalized patients who tested negative for SARS-CoV-2 and without ARI signs and symptoms | ≥14 days before reference date (date of hospitalization for cases) | (1–OR) X 100<br>OR: Prior vaccination in cases vs. controls | Logistic regression |
| Tenforde MW, et al. [19] | • VE: Odds of prior SARS-CoV-2 vaccination in hospitalized cases with COVID-19 (symptoms and a positive RT-PCR/clinical test) vs. controls | • Test-negative controls: Hospitalized patients who tested negative for SARS-CoV-2, but had COVID-19-like illness<br>• COVID-19-like illness-negative controls: Hospitalized patients who tested negative for SARS-CoV-2 and did not have COVID-19-like illness | • $\geq 14$ days before reference date (date of hospitalization for cases)<br>• Full surveillance period and separately for March–May and June–July 2021, because of increased circulation of Delta variant in the US | (1–OR) X 100<br>OR: Prior vaccination in cases vs. controls | Logistic regression |
| Khan N & Mahmud N. [20] | • VE: SARS-CoV-2 infection (positive RT-PCR test)<br>• VE: Severe COVID-19 (hospitalization or death)<br>• VE: All-cause mortality (extracted from vital status file in VHA dataset) | Unvaccinated controls | • $\geq 7$ days after 2<sup>nd</sup> dose<br>• Median fully vaccinated: 38 days<br>• Median unvaccinated: 123 days | (1 – incidence vaccinated/incidence unvaccinated) X 100 | • Inverse probability weighted (IPW) adjusted models<br>• Cox proportional hazards regression |
| Polinski JM, et al. [21] | • VE: SARS-CoV-2 infection (medical claim with COVID-19 diagnosis code and/or a positive RT-PCR test)<br>• VE: COVID-19-related hospitalization (discharge diagnosis of COVID-19 or a | Unvaccinated controls | • $\geq 14$ days after single dose up to 5 months | (1–HR) X 100<br>HR: COVID-19 infection/hospitalization in vaccinated vs. unvaccinated | • Propensity score matching<br>• Incidence rates calculated per 1,000 person-years<br>• Cox regression models used to |
|  | recorded infection within 21 days before admission) |  |  |  | calculate hazard ratios<br>• VE estimates were corrected for under-recording in data source |
| Dagan N, et al.;<br>Barda N, et al.<br>[22,23] | • VE: SARS-CoV-2 infection (positive RT-PCR test)<br>• VE: Symptomatic COVID-19 (documented in EMR)<br>• VE: Severe COVID-19 illness (according to NIH criteria [31])<br>• VE: COVID-19-related hospitalization (admission records)<br>• VE: COVID-19-related death (admission records); not assessed in IC | Matched unvaccinated controls | 7 days after 2<sup>nd</sup> dose up to 28 days: mean follow-up: 18 days in subgroup analyses | 1 - RR<br>RR: vaccinated vs. unvaccinated | • Matching analysis<br>• Kaplan-Meier analysis to compute risk of outcomes and then calculated risk ratios |
| Chodick G, et al.<br>[24] | • VE: SARS-CoV-2 infection ($\geq 1$ positive RT-PCR test)<br>• VE: Symptomatic COVID-19 (documented in EMRs)<br>• VE: Risk of COVID-19-related hospitalization/death among infected (extracted from EMRs); VE not reported in IC | Between 7 to 27 days after 2<sup>nd</sup> dose (protection-period) vs. days 1 to 7 after the 1<sup>st</sup> dose (reference period) | Days 7-27 after 2<sup>nd</sup> dose vs. days 1-7 after 1<sup>st</sup> dose | (1 – Relative Risk) X 100<br>Relative Risk: Protection-period vs. reference-period | • Logistic regression was used to estimate odds ratios for symptomatic COVID-19 among infected cases<br>• Cox proportional hazards model were used to calculate adjusted hazard ratios for COVID-19-related hospitalization/death. |
| Yelin I, et al. [25] | • VE: SARS-CoV-2 infection (positive RT-PCR test)<br>• VE: Symptomatic COVID-19; not assessed in IC | Unvaccinated controls | • 1-11 days<br>• 12-28 days<br>• 29-50 days (IC)<br>All post-vaccination | (1-OR) X 100<br>OR: Positive test/symptomatic infection in vaccinated vs. unvaccinated | Logistic regression |
| Whitaker HJ, et al. [26] | • VE: Symptomatic COVID-19 (diagnosis of COVID-19 or clinical illness consistent with COVID-19 within 10 days before or after a positive RT-PCR test) | Test-negative controls: Patients with symptoms within 10 days of SARS-CoV-2 negative test | ≥14 days after 2<sup>nd</sup> dose up to ~6.5 months | Adjusted VE OR<br>OR: Cases in vaccinated vs. unvaccinated | Logistic regression |
| Chemaitelly H, et al. [27] | • VE: SARS-CoV-2 infection (positive RT-PCR test)<br>• VE: Severe COVID-19 illness (according to WHO criteria [32]) | Unvaccinated controls | • ≥14 days after 2<sup>nd</sup> dose<br>• ≥42 days after 2<sup>nd</sup> dose<br>• ≥56 days after 2<sup>nd</sup> dose | 1 - HR<br>HR: vaccinated vs. unvaccinated | A proportional hazards model was used to calculate adjusted hazard ratios |

COVID-19: Coronavirus disease 2019; EMR: Electronic medical records; HR: Hazard ratio; IC: Immunocompromised; NIH: National Institutes of Health; OR: Odds ratio; RR: Risk ratio; RT-PCR: Reverse transcription polymerase chain reaction; SARS-CoV-2: Severe acute respiratory syndrome coronavirus 2; VE: Vaccine effectiveness; VHA: Veterans Health Administration; WHO: World Health Organization

**Table 5.**
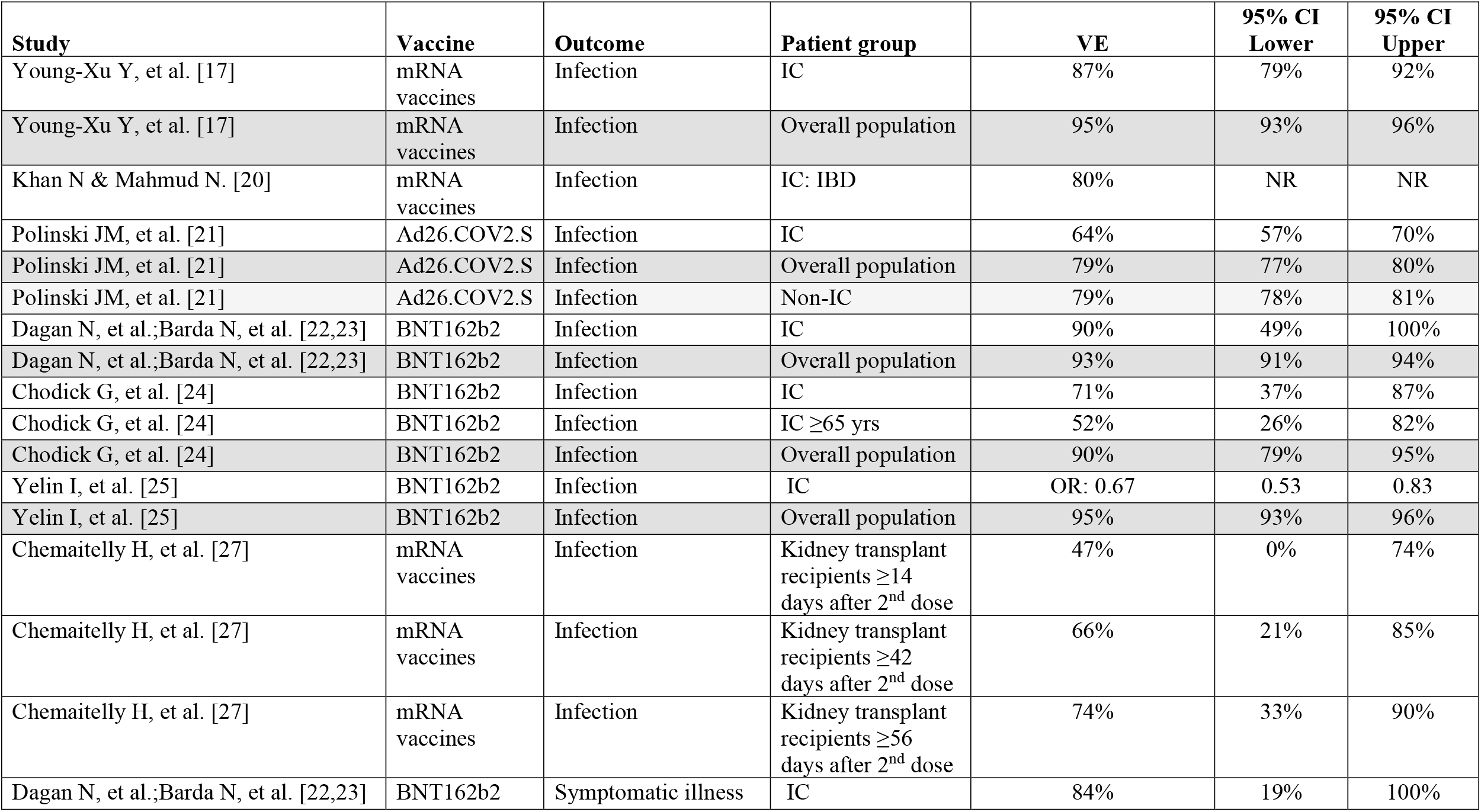
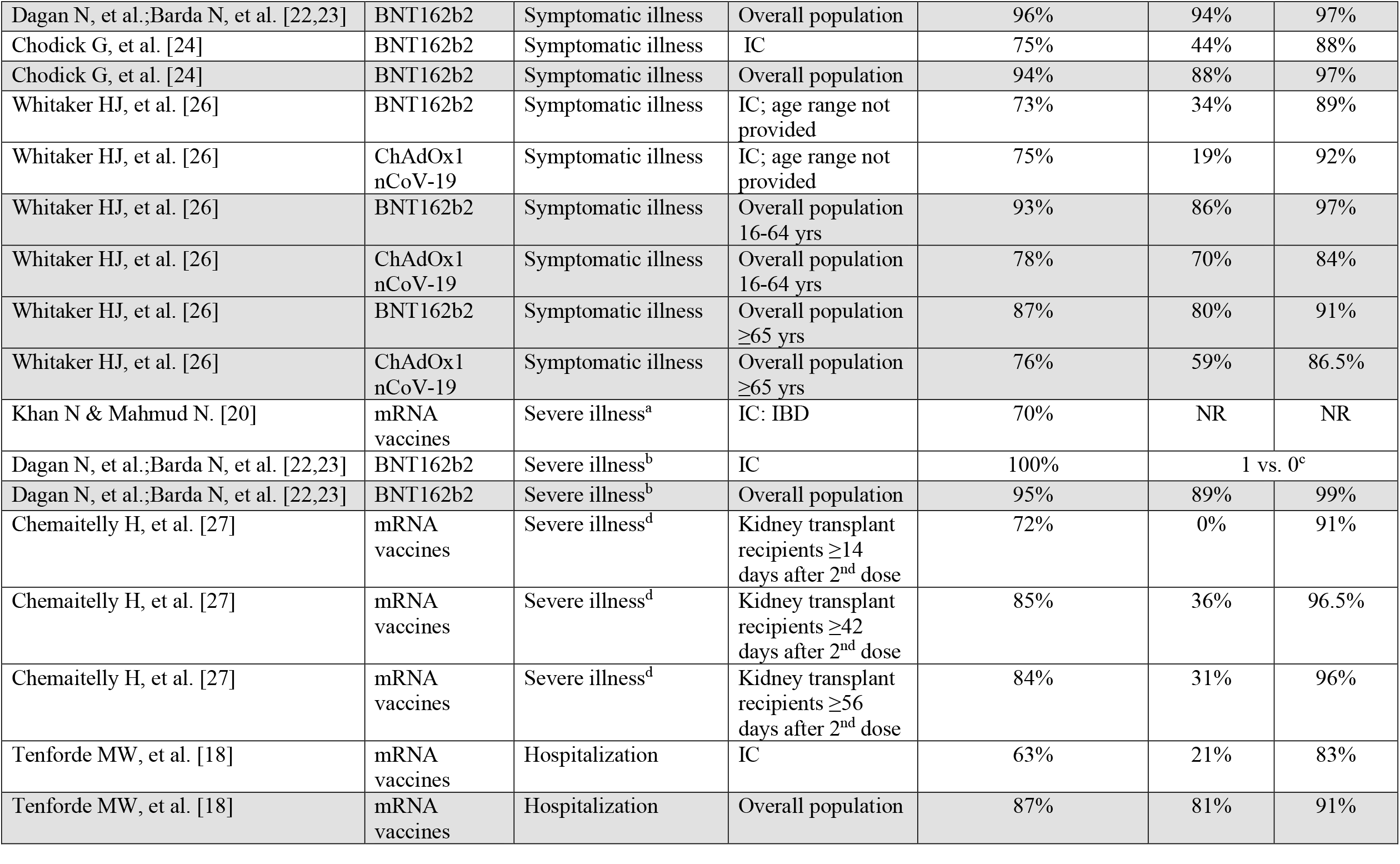
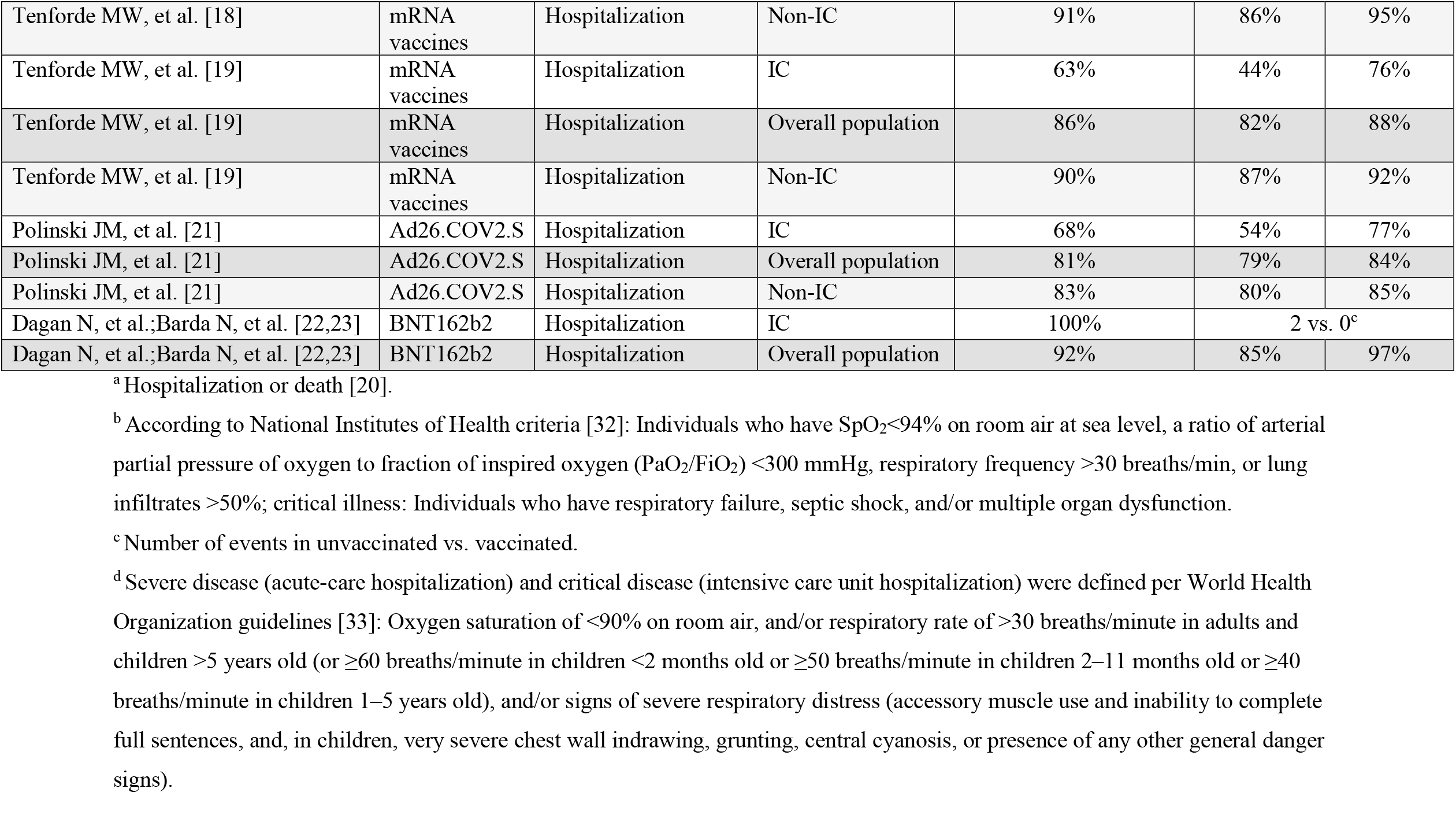
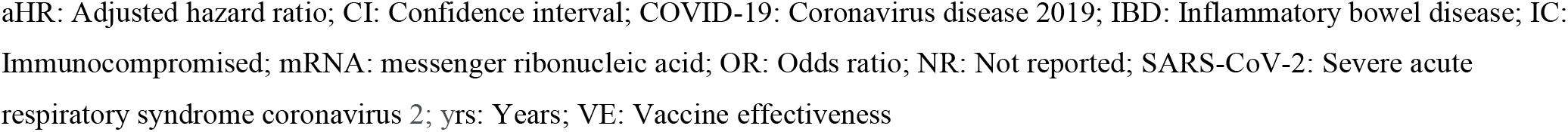
Vaccine effectiveness against SARS-CoV-2 infection, symptomatic COVID-19 illness, severe COVID-19 illness, and COVID-19-related hospitalization across studies.

**Figure 1.**
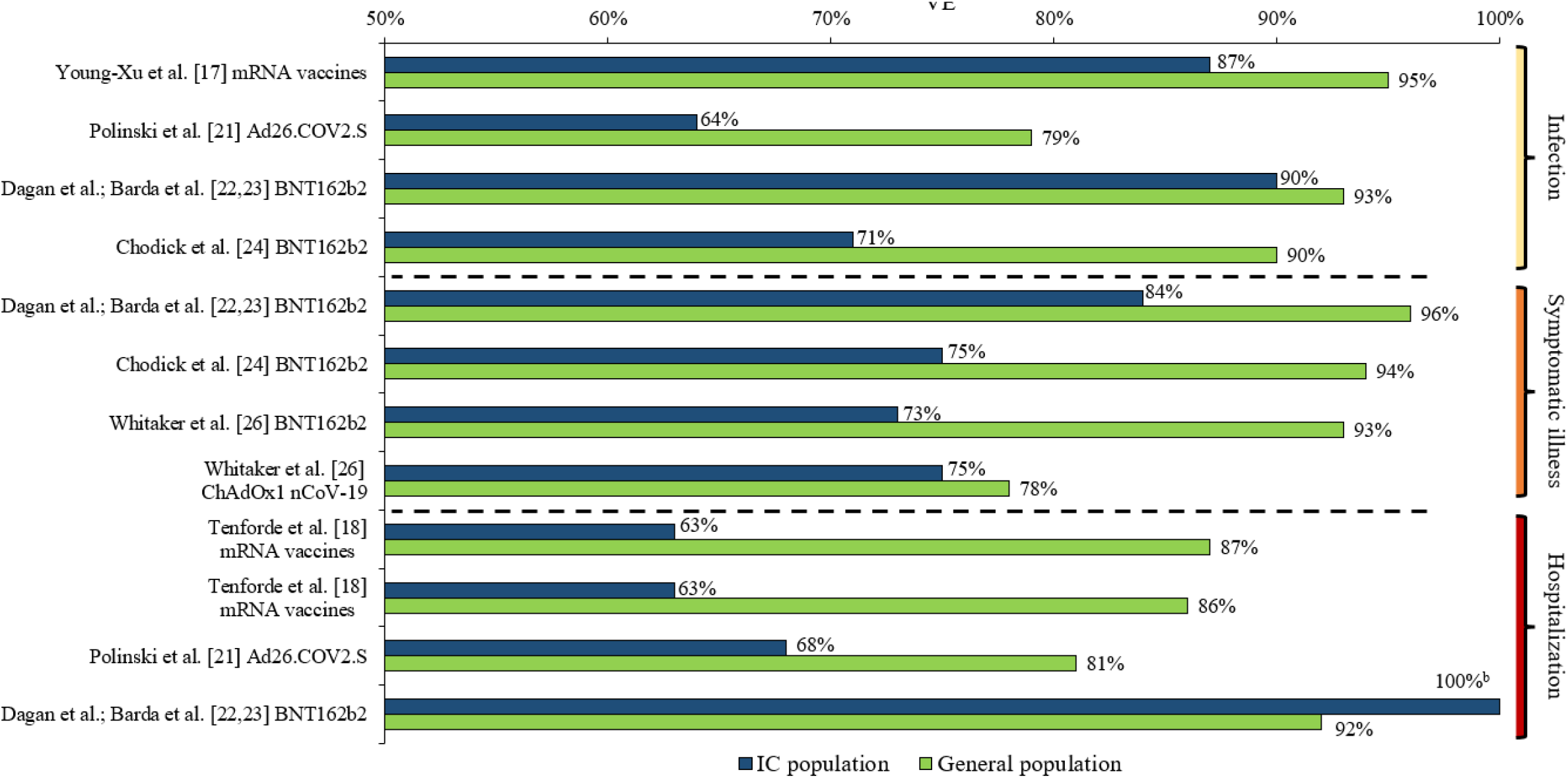
VE against SARS-CoV-2 infection, symptomatic COVID-19 illness, and COVID-19-related hospitalization in IC populations versus general populations^a^. ^a^ See Table 5 for VE including 95% CIs. ^b^ Dagan et al.; Barda et al. [22,23] reported a BNT162b2 VE against COVID-19-related hospitalization of 100% in their IC population; however, only two such events occurred in the unvaccinated IC group and none in the vaccinated group. CI: Confidence interval; COVID-19: Coronavirus disease 2019; IC: Immunocompromised; SARS-CoV-2: Severe acute respiratory syndrome coronavirus 2; VE: Vaccine effectiveness

#### 3.3.1 COVID-19 VE against SARS-CoV-2 infection

Seven studies (Young-Xu et al. [17]; Khan & Mahmud [20]; Polinski et al. [21]; Dagan et al.; Barda et al. [22,23]; Chodick et al. [24]; Yelin, et al. [25]; and Chemaitelly et al. [27]) assessed VE against SARS-CoV-2 infection in IC populations; the time periods of measured VE varied in these seven studies (e.g, ≥7 or ≥14 days after second vaccine dose to end of follow-up; 5 month maximum). Additionally, the definition of SARS-CoV-2 infection differed to some extent. All studies required a positive RT-PCR test; Young-Xu et al. [17] also included a positive antigen test, while Polinski et al. [21] defined SARS-CoV-2 infection as a medical claim with a COVID-19 diagnosis code (85% of cases) and/or a positive RT-PCR test (15% of cases). Across the five studies that assessed VE of mRNA vaccines against SARS-CoV-2 infection [17,20,22,23,24,25], VE ranged from 52% to 90% in the IC populations, versus a VE of 90% to 95% in the overall study populations. The lowest mRNA VE (52%; 95% CI: 26%-82%) was observed among individuals who were ≥65 years of age and IC (IC conditions included: hematopoietic cell or solid organs transplant, immunosuppressive medication usage, asplenia, and chronic renal failure [advanced kidney disease, dialysis, or nephrotic syndrome]) [24]. In this study by Chodick et al. [24], mRNA VE was 71% (95% CI: 37%-87%) among those who were IC (age range not provided). The highest VE against SARS-CoV-2 infection (90%; 95% CI: 49%-100%) among an IC population was observed in the Israeli study of Dagan et al. [22,23] (IC conditions included HIV, asymptomatic HIV, organ transplant, bone marrow or hematopoietic stem cell transplant, immunosuppressive medication usage). Young-Xu et al. [17], additionally conducted a post-hoc IC subgroup analysis of mRNA VE against SARS-CoV-2 infection in US veterans with hematological malignant neoplasms, which was 69% (95% CI: 17%-88%).

In the study of Chemaitelly et al. [27] of kidney transplant recipients, mRNA VE was measured at different time periods post-second dose, and as the duration increased, VE against SARS-CoV-2 infection increased from 47% (95% CI: 0%-74%) at ≥14 days to 66% (95% CI: 21%-85%) and 74% (95% CI: 33%-90%) at ≥42 days and ≥56 days, respectively, indicating vaccine protection in this IC patient group did not reach a high level until several weeks after the second dose. Yelin et al. [25] assessed VE of BNT162b2 and only reported an odds ratio (0.67; 95% CI: 0.53-0.83), and not VE for the IC population, relative to the overall population with a VE of 95% (95% CI: 93%-96%), indicating that the IC population had a 33% reduction in VE relative to the overall population. Polinski et al. [21] assessed Ad26.COV2.S VE against SARS-CoV-2 infection, which was 64% (95% CI: 57%-70%) in the IC population and 79% in both the overall (95% CI: 77%-80%) and non-IC (95% CI: 78%-81%) populations.

#### 3.3.2. VE against symptomatic COVID-19 illness

Three studies (Dagan et al.; Barda et al. [22,23]; Chodick et al. [24]; and Whitaker et al. [26]) assessed VE against symptomatic COVID-19 illness in IC populations; Dagan et al.; Barda et al. [22,23] assessed VE 7-28 days after the second vaccine dose, Chodick et al. [24] assessed VE on days 7-27 after the second vaccine dose versus days 1-7 after the first vaccine dose, and Whitaker et al. [26] assessed VE ≥14 days after the second vaccine dose up to approximately 6.5 months. The definition of symptomatic COVID-19 illness was generally similar across the studies; Dagan et al. [22]; Barda et al. [23] and Chodick et al. [24] required documentation of symptomatic COVID-19 illness in EMRs, while Whitaker et al. [26] required a diagnosis of COVID-19 or clinical illness consistent with COVID-19 within 10 days before or after a positive RT-PCR test. In the two Israeli studies by Dagan et al.; Barda et al. [22,23] and Chodick et al. [24], BNT162b2 VE against symptomatic COVID-19 illness was 84% (95% CI: 19%-100%) and 75% (95% CI: 44%-88%) in the IC populations, respectively, while it ranged 94% (95% CI: 88%-97% [24]) to 96% (95% CI: 94%-97% [22,23]) in the overall study populations. From the study conducted in England, Whitaker et al. [26] reported a BNT162b2 VE of 73% (95% CI: 34%-89%) against symptomatic COVID-19 illness among the IC population (age range not provided); VE in the overall population 16-64 years of age was 93% (95% CI: 86%-97%), while it was 87% (95% CI: 80%-90%) for those ≥65 years. Whitaker et al. [26] also assessed ChAdOx1 nCoV-19 VE against symptomatic COVID-19 illness, which was 75% (95% CI: 19%-92%) among the IC population, 78% (95% CI:70%-84%) among the overall population 16-64 years of age, and 76% (95% CI: 59%-86.5%) among the overall population ≥65 years of age.

#### 3.3.3 VE against severe COVID-19 illness

Three studies (Khan & Mahmud [20]; Dagan et al.; Barda et al. [22,23], and Chemaitelly et al. [27]) assessed mRNA VE in IC populations against severe COVID-19 illness, which was defined differently across these studies. Khan & Mahmud [20] defined severe COVID-19 illness as hospitalization or death, while Dagan et al.; Barda et al. [22,23] defined it according to National Institutes of Health (NIH) criteria [31], and Chemaitelly et al. defined it as per the World Health Organization (WHO) criteria [32]. Khan & Mahmud [20] reported a mRNA VE against severe illness of 70% (95% CI: NR) among their IBD patient population. Dagan et al.; Barda et al. [22,23] reported a BNT162b2 VE of 100% (95% CI: could not be determined) against severe illness in the IC population compared to 95% (95% CI: 89%-99%) in the overall population; however, the interpretation of such a finding should consider the low statistical power due to the small numbers of events (i.e., only one such event of severe illness occurred in the unvaccinated IC group and none in the vaccinated group). Similar to VE against SARS-CoV-2 infection, Chemaitelly et al. [27] observed an increase in mRNA VE against severe illness with increased time post-second dose in kidney transplant recipients, from 72% (95% CI: 0%-91%) at ≥14 days to 85% (95% CI: 36%-96.5%) and 84% (95% CI: 31%-96%) at ≥42 days and ≥56 days, respectively.

#### 3.3.4. VE against COVID-19-related hospitalization

Four studies assessed VE against COVID-19-related hospitalization in IC populations; three (Tenforde et al. [18]; Tenforde et al. [19]; Dagan et al.; Barda et al. [22,23]) assessed VE of mRNA vaccines and Polinski et al. [21] assessed VE of Ad26.COV2.S. In the first CDC study by Tenforde et al. [18], mRNA VE against COVID-19-related hospitalization was 63% (95% CI: 21%-83%) in the IC population, 87% (95% CI: 81%-91%) in the overall study population, and 91% (95% CI: 86%-95%) in the non-IC population. The second CDC study by Tenforde et al. [19], which included nearly three times more hospitalized patients, had similar findings, with mRNA VE against COVID-19 related hospitalization reported at 63% (95% CI: 44%-76%) in the IC population, 86% (95% CI: 82%-88%) in the overall study population, and 90% (95% CI: 87%-92%) in the non-IC population over the full surveillance period (March-July 2021). Although overall VE in the IC population was lower than that in the non-IC population, it was sustained over the two study periods (March-May: 2-12 weeks and June-July: 13-24 weeks post full vaccination), which was consistent to the sustained VE observed in the overall population [19]. Dagan et al.; Barda et al. [22,23] reported a BNT162b2 VE against COVID-19-related hospitalization of 100% (95% CI: could not be determined) in their IC population; however, only two such events occurred in the unvaccinated IC group and none in the vaccinated group; VE was 92% (95% CI: 85%-97%) in the overall study population. In the study of Polinski et al. [21], VE of Ad26.COV2.S against COVID-19-related hospitalization was 68% (95% CI: 54%-77%) in the IC population compared to 81% (95% CI: 79%-84%) in the overall study population and 83% (95% CI: 80%-85%) in the non-IC population [21].

## 4. Discussion

This targeted literature review of 10 real-world studies conducted in four different countries gives an early view of COVID-19 VE in IC populations. Among the fully vaccinated IC populations included in the studies, VE of widely available COVID-19 vaccines ranged from 64% to 90% against SARS-CoV-2 infection, 73% to 84% against symptomatic COVID-19 illness, 70% to 100% against severe COVID-19 illness, and 63% to 100% against COVID-19-related hospitalization. COVID-19 VE for most outcomes in the IC populations included in these studies was lower than in the general populations, in which VE ranged from 79% to 95% against SARS-CoV-2 infection, from 76% to 96% against symptomatic COVID-19 illness, and from 81% to 92% against COVID-19-related hospitalization. Important to consider when interpreting the reported VE estimates for the IC populations are the accompanying confidence intervals, ranges of which were wider than those reported among the general populations across studies; such findings are related to the smaller sample sizes of the IC populations, but also stress the variability in COVID-19 VE across individuals with various IC conditions within overall IC populations. Moreover, the confidence intervals ranged substantially even among kidney transplant recipients only in the study of Chemaitelly et al. [27] suggesting that even when COVID-19 VE is assessed in one specific IC patient group, there is significant variability among individuals. These summarized findings provide a preliminary evidence base supporting greater protective measures to prevent COVID-19 infection and associated illness in those who are IC.

In the rapidly changing COVID research environment, new studies continue to be conducted, published or posted as preprints. Some of these noteworthy, late-breaking studies were not available during the study timeframe as defined for this current review. A recently medRxiv posted systematic review and meta-analysis of 54 observational and longitudinal studies of general populations [33], wherein COVID-19 VE was assessed among fully vaccinated (post-second vaccine dose) individuals, estimated a pooled VE for the BNT162b2, mRNA-1273, and ChAdOx1 nCoV-19 vaccines against SARS-CoV-2 infection of 87% (pooled odds ratio [OR] = 0.13; 95% CI: 0.08-0.21). Against COVID-19 related hospitalization, a pooled VE of 89% (pooled OR = 0.11; 95% CI: 0.07-0.17) was estimated [33]. The findings of this meta-analysis of COVID-19 VE are relatively consistent and in the range of the COVID-19 VE estimates reported in the general populations of the summarized studies herein. Altogether, these study findings emphasize the effectiveness of widely used COVID-19 vaccines across populations from different countries.

Embi et al. [34] published a study on November 5, 2021, in which COVID-19 mRNA VE against COVID-19-related hospitalization was estimated in a fully vaccinated (i.e., after completing 2 doses of an mRNA vaccine with ≥14 days prior to index hospitalization date) US population. This test-negative designed study utilized data from the VISION network, a CDC collaboration with seven US healthcare systems and research centers, including 187 hospitals in nine US states; it included over 89,000 COVID-19-associated hospitalizations of IC and immunocompetent adults [34]. The IC population in this study was defined as individuals with a diagnosis of solid malignancy, hematologic malignancy, rheumatologic or other inflammatory disorders, other intrinsic immune conditions or immunodeficiencies, or organ or stem cell transplants; immunosuppressive medication usage was not included in this study since the data were not available [34]. Embi et al. [34] reported a COVID-19 mRNA VE against COVID-19-associated hospitalization of 77% (95% CI: 74%-80%) among 10,564 fully vaccinated IC individuals during January 17 through September 5, 2021, [34]and a VE of 90% (95% CI: 89%-91%) among those considered immunocompetent [34]. Additionally, Embi et al. [34] assessed COVID-19 mRNA VE before and during Delta variant predominance in the US; they consistently found a lower VE against COVID-19-associated hospitalization among the IC compared to the immunocompetent before (76%; 95% CI: 69%-81% versus 91%; 95% CI: 90%-93%) and during Delta variant predominance (79%; 95% CI: 74%-83% versus 90%; 95% CI: 89%-91%). COVID-19 mRNA VE in the IC population relative to the immunocompetent population did not significantly differ by age group (18-64 years of age and aged ≥65 years) or mRNA vaccine type, nor by time periods of assessment [34].

In the four studies reviewed herein that estimated VE against COVID-19-related hospitalization, VE ranged from 63% to 100% in the IC populations and 81% to 92% in the general populations [18,19,21,22,23]. Only Tenforde et al. [19] included a time period in which the Delta variant emerged as predominant; similar to the above findings of Embi et al. [34], during emerging Delta variant predominance (June-July 2021), COVID-19 mRNA VE against COVID-19-associated hospitalization did not significantly change among the IC or the overall study population from the earlier study period of March-May 2021. Embi et al. [34] also performed subgroup analyses among the IC population, in which mRNA VE against COVID-19-related hospitalization was estimated between January 17 and September 5, 2021 in organ or stem cell transplant recipients at 59% (95% CI: 38%-73%), in those with solid malignancy at 79% (95% CI: 73%-84%), in those with hematologic malignancy at 74% (95% CI: 62%-83%), in those with intrinsic immune conditions or primary immunodeficiencies at 73% (95% CI: 66%-80%), and in those with rheumatic or inflammatory disorders at 81% (95% CI: 75%-86%); all IC subgroups exhibited lower VE than among the immunocompetent population of this study. Of the summarized studies in this review, only Tenforde et al. [18] reported mRNA VE against COVID-19 related hospitalization for a subgroup of the IC population; the estimated VE was 51% (95% CI: -31%-82%) against COVID-19 related hospitalization for IC patients with an active solid organ or hematologic malignancy or solid organ transplant. These study findings further highlight that certain IC patient groups exhibit significantly lower COVID-19 VE than the general population, as well as the variability in VE between groups with different IC conditions, further warranting greater research of VE in particular IC groups.

On November 17, 2021, Galmiche et al. [35] published a systematic review of studies, in which COVID-19 VE in IC populations in real-world settings was assessed in four of the included studies. The other studies included in this systematic review assessed COVID-19 vaccine immunogenicity in IC populations (N=157 studies) and one study assessed vaccine efficacy in a clinical trial setting [35]. Three of the four studies that assessed COVID-19 VE in IC populations included in this systematic review, Tenforde et al. [18], Khan & Mahmud [20], and Chodick et al. [24], have already been included in our targeted literature review. The fourth study by Aslam et al. [36], reported incidence rates of symptomatic COVID-19 illness in solid organ transplant recipients (N=2,151) and not a calculated VE; in those who were vaccinated the incidence rate was 0.065 per 1000/person days (95% CI: 0.024-0.17) and in those who were unvaccinated or partially vaccinated, the incidence rate was 0.34 per 1000/person days (95% CI: 0.26-0.44).

In a US real-world study of nearly 1.2 million people fully vaccinated with the BNT162b2 mRNA vaccine, over 212,000 (18%) individuals were designated as having an IC condition [37]. This study utilized the broadest IC case algorithm of real-world studies to date, wherein 12 mutually exclusive IC conditions were identified (e.g., symptomatic HIV, solid/hematologic malignancy, organ transplant, rheumatologic/inflammatory condition, primary immunodeficiency, chronic kidney disease, usage of immunosuppressive/antimetabolite medication) [37]. Although this study did not directly measure VE, it reported the number of COVID-19 vaccine breakthrough infections following a second BNT162b2 dose between December 10, 2020 and July 8^th^, 2021 [37]. The total number of breakthrough infections was low (N=978; 0.08%) but nearly 40% of cases occurred among the IC population, which only accounted for approximately 18% of the overall study population [37]. The calculated incidence rate of COVID-19 vaccine breakthrough infections was 2.6 times higher among the IC population than in the non-IC population (0.89 vs. 0.34 per 100 person-years) [37]. Moreover, approximately 60% (N=74 of 124) of the breakthrough infections that resulted in hospitalization and 100% (N=2 of 2) of those that resulted in inpatient death, occurred in the IC population [37]. In this study, subgroup analyses of the 12 IC condition groups were also conducted; organ transplant recipients excluding bone marrow transplant had the highest incidence rate of breakthrough infections (3.66 per 100 person-years) [37]. Additionally, compared to the incidence rate among the overall IC population in this study, incidence rates of breakthrough infections were higher in those who had >1 IC condition, those with usage of antimetabolites, those with a primary immunodeficiency, those with a hematologic malignancy, and those with kidney disease [37]. The findings of this study underscore the need to standardize the definition of IC across research studies evaluating COVID-19 VE and to also conduct studies of specific IC patient groups, so that a risk stratification can be established across the overall IC population.

At the time this review was written, only 10 real-world studies, four of which were preprints without peer-review, were available that assessed COVID-19 VE in IC populations. Although our approach of including preprints for this targeted literature review strengthens the comprehensiveness of this review, we acknowledge the potential limitations in the reproducibility of this review and the quality of the collected evidence base. Moreover, of the 10 included studies, study designs, follow-up periods after full vaccination, IC definitions and IC populations, methods of computing VE, and adjustment for confounders significantly varied across these real-world studies. Hence, a comparison of study findings or a meta-analysis estimating the pooled VE for outcomes of interest was considered unfeasible. As discussed earlier, the most notable inconsistency across the studies summarized in this review, was the substantial variability in the definitions of IC populations. In this context, the COVID-19 VE estimates across these studies should be interpreted cautiously.

Additionally, the reviewed studies had limited follow-up after vaccination ranging from 7 days to 6.5 months. Four studies by Tenforde et al. [19], Polinski et al. [21], Whitaker et al. [26], and Chemaitelly et al. [27] included VE analyses during time periods of Delta variant predominance; however, only Tenforde et al. [19] reported, albeit in a figure only, mRNA VE in the IC during March to May (Alpha variant predominance) and June to July (Delta variant emerging as predominant) 2021. The study period in Tenforde et al. [19] went through July 2021, which covered only the early period of Delta variant predominance in the US (approximately the first six weeks), and a Delta-specific VE was not reported [19]. As mentioned earlier, Embi et al. [34] did not observe a significant change in COVID-19 mRNA VE against COVID-19-associated hospitalization among IC or immunocompetent individuals during Delta variant predominance compared with an earlier time period. Altogether, the studies summarized in this review covered up to eight months after COVID-19 vaccines became available. Thus, waning COVID-19 vaccine protection remains relatively undescribed, particularly among the IC, and further follow-up studies are needed to better understand not only waning vaccine protection but also the impact of increased vaccine protection with an additional dose. Only a few studies performed subgroup analyses by IC condition groups or severity of IC conditions. The included studies were also from only four countries including the US, Israel, England, and Qatar. Therefore, study findings may not be generalizable to IC population in other countries, especially in countries where particular IC conditions are endemic to the region. While this review provides an early view of COVID-19 VE in IC populations, mostly as an aggregate group, further study is warranted.

## 5. Expert opinion

As the COVID-19 pandemic continues across the world and if in the future, COVID-19 becomes endemic to societies, it may be of clinical utility to more consistently and precisely define IC populations across research studies evaluating COVID-19 VE. A consensus on defining IC condition groups will provide more useful evidence for policymakers and healthcare providers in the decision-making process when recommending and updating vaccination protocols and treating patients at high-risk for COVID-19. Across the studies included in this review, only two IC conditions, organ transplant and immunosuppressive medication usage, were common in the definitions of IC populations. Only a few studies included in this review focused on particular IC conditions and only one study included CKD as an IC condition. A consensus on the list of immunosuppressive medications to designate individuals as IC also needs to be developed. Moreover, it may be useful to stratify overall IC populations into low-, medium-, and high-risk patient groups for COVID-19 illness. This may also involve the identification of IC groups with comorbidities known to increase the risk for severe COVID-19 (e.g., older age, type 2 diabetes, obesity) [38,39] and their risk stratification. Furthermore, individuals with IC conditions that are endemic to certain countries and regions that heighten the risk for COVID-19 illness may also need to be identified so that the necessary preventive and protective measures can be put in place.

This review highlights the most current findings of real-world studies that have assessed COVID-19 VE in IC populations. Our summarized findings provide preliminary evidence that individuals who are IC require greater protective measures to prevent COVID-19 infection and associated illness; hence, should be prioritized while implementing recommendations of additional COVID-19 vaccine doses. Indeed, in the US, the CDC recommends an additional primary mRNA COVID-19 vaccine dose for moderately or severely IC people at least 12 years of age who received a two-dose mRNA vaccine primary series [40], and countries, including Israel, the United Kingdom, and France, as well as the World Health Organization, have similar recommendations [41,42]. In a recent study of approximately 22,000 US survey respondents with comorbid conditions all participating in an online health community, of whom 27% reported having cancer and 23% reported having an autoimmune disease, approximately 20% expressed COVID-19 vaccine hesitancy [43]. In light of this finding, in addition to the substantial number of people who have IC conditions and/or take immunosuppressive medications, and the potential for waning COVID-19 VE and emergence of new SARS-CoV-2 variants, it is critical to rapidly advance our understanding of COVID-19 VE and duration of response among IC populations, including specific IC condition groups and IC individuals who have other COVID-19 risk factors (e.g., elderly, comorbidities, etc.), as the COVID-19 pandemic continues worldwide. The importance of this undertaking is explicitly emphasized by the recent emergence of the highly transmissible Omicron variant, which again underscores the ongoing need to provide the most up to date scientific information to decision makers so that measures, such as immunization scheduling and additional dose/booster prioritization, can be rapidly implemented.

## Data Availability

Data availability statement: All data contained within this review were extracted from the published/preprint referenced articles and are available to the public.

https://pubmed.ncbi.nlm.nih.gov/

https://www.medrxiv.org/

https://khub.net/

## Article highlights

- Scientific evidence gained from real-world studies conducted in multiple countries is increasingly showing that widely available COVID-19 vaccines, including BNT162b2 (Pfizer/BioNTech), mRNA-1273 (Moderna), Ad26.COV2.S (Janssen), and ChAdOx1 nCoV-19 (Oxford/AstraZeneca), are highly effective for protecting against SARS-CoV-2 infection, symptomatic COVID-19 illness, and COVID-19-related hospitalization and death.
- From July through October of 2021, several countries across the world issued recommendations for increased COVID-19 vaccine protection for individuals with one or more immunocompromised (IC) conditions.
- As new COVID-19 vaccine recommendations are implemented and updated over time in response to the evolving COVID-19 pandemic, it is necessary to rapidly and more comprehensively understand the effectiveness of COVID-19 vaccines in IC populations.
- In this review, we have summarized the findings of real-world studies that have assessed COVID-19 VE in IC populations.
- Among the fully vaccinated IC populations included in the reviewed studies, VE of widely available COVID-19 vaccines ranged from 64% to 90% against SARS-CoV-2 infection, 73% to 84% against symptomatic COVID-19 illness, 70% to 100% against severe COVID-19 illness, and 63% to 100% against COVID-19-related hospitalization.
- VE for most outcomes in the IC populations included in these studies was lower than in the general populations, in which VE ranged from 79% to 95% against SARS-CoV-2 infection, from 76% to 96% against symptomatic COVID-19 illness, and from 81% to 92% against COVID-19-related hospitalization.
- Our summarized findings provide preliminary evidence that individuals who are IC require greater protective measures to prevent COVID-19 infection and associated illness; hence, should be prioritized while implementing recommendations of additional COVID-19 vaccine doses.

## Authorship contributions

All named authors meet the International Committee of Medical Journal Editors (ICMJE) criteria for authorship for this article. All authors contributed to study conception and design, analysis, and interpretation, drafting and revising of the manuscript, and have given their approval for this manuscript version to be published.

## Data availability statement

All data contained within this review were extracted from the published/preprint referenced articles and are available to the public.

